# Measurement lessons of a repeated cross-sectional household food insecurity survey during the COVID-19 pandemic in Mexico

**DOI:** 10.1101/2020.08.04.20167650

**Authors:** P. Gaitán-Rossi, M. Vilar-Compte, G. Teruel, R. Pérez-Escamilla

## Abstract

**Objective:** To validate the telephone modality of the Latin American and Caribbean Food Security Scale (ELCSA) included in three waves of a phone survey to estimate the monthly household food insecurity (HFI) prevalence during the COVID-19 pandemic in Mexico.

**Design:** We examined the reliability and internal validity of the ELCSA scale in three repeated waves of a cross-sectional surveys with Rasch models. We estimated the monthly prevalence of food insecurity in the general population and in households with and without children, and compared them with a national 2018 survey. We tested concurrent validity by testing associations of HFI with socioeconomic status and anxiety.

**Setting:** ENCOVID-19 is a monthly telephone cross-sectional survey collecting information on the well-being of Mexican households during the pandemic lockdown. Surveys used probabilistic samples and we used data from April (n=833), May (n=850), and June 2020 (n=1,674).

**Participants:** Mexicans 18 years or older who had a mobile telephone.

**Results:** ELCSA had adequate model fit and HFI was associated, within each wave, with more poverty and anxiety. The COVID-19 lockdown was associated with an important reduction in food security; decreasing stepwise from 38.9% in 2018 to 24.9% in June 2020 in households with children.

**Conclusions:** Telephone surveys are a feasible strategy to monitor food insecurity with ELCSA

## Introduction

The number of people with severe food insecurity has been rising globally since 2014 and the COVID-19 pandemic will likely reduce food security even further (1). Several socioeconomic and health pathways can contribute to such reduction. Even though the association between household income and food security is well established (2), the COVID-19 pandemic constitutes an important external socioeconomic shock to households because its effects in unemployment, poverty, and a subsequent reduction in food purchases (3). The shock might strike harder in households that were already vulnerable prior to the pandemic. In the United States, households that allocate a higher income share on food are affected the most by economic shocks (4) and previous studies have found that severe economic crises, such as the one in 2008, reduced food security in Mexican households and had a larger effect among households with lower income (5). The current pandemic crisis has been long lasting, and also affecting the food security of households that were not poor prior to the pandemic due to debt, temporary or permanent job loss, or catastrophic illness (8). In addition to its negative economic impacts, the needed social distancing measures are disrupting food systems leading to increased food prices and making it more difficult to access healthy foods (6). Moreover, these measures may hinder the access to food assistance programs, such as school meals (7). A serious concern is that it may also affect obesity risk and health outcomes by increasing the consumption of ultra-processed foods, sedentarism, and by reducing access to health services (3).

Another consequence of experiencing food insecurity is poor mental health. A global analysis of the effects of food insecurity on mental health status found a consistent dose-response; increasing food insecurity amplified diverse psychosocial stressors that were linked to heightened anxiety (8). A recent meta-analysis identified that food insecurity has a significant effect on the likelihood of being stressed or depressed (9). Food security in households with children requires particular attention (10) because it has been associated with developmental risk and, in low and middle-income countries, with lower vocabulary skills (11). These child development delays can be mediated by depression or anxiety in caregivers (12). The disruption of the COVID-19 pandemic can amplify these adverse outcomes because the required attention may be insufficient in fragile mental health care systems in low and middle income countries (13). The COVID-19 pandemic is therefore a multi-causal and complex syndemic that we hypothesize wil worsen food insecurity (3).

Experienced-based food security scales –such as the Escala Latinoamericana y Caribeña de Seguridad Alimentaria (ELCSA) (14)– have been shown to be cost-effective and valid to assess food insecurity via face-to-face interviews in most countries in Latin America, Africa, and Asia (15). The COVID-19 pandemic made it impossible to conduct much needed HFI monitoring via face-to-face surveys in Mexico. Hence a repeated, low-cost remote HFI assessment was warranted (16). Telephone surveys were considered as a feasible strategy to address this need as interview mode (in-person vs telephone) has been shown before to have small effects on food security prevalence estimates; the adjusted odds ratio of food insecurity comparing in-person with telephone interview in the United States was 1.036 (P= 0.088) (17). Moreover, experienced-based food security scales had already been applied by telephone in middle- and high-income countries with at least 80% telephone coverage (15). To the extent of our knowledge, however, the assessment of HFI with an experienced-based food security scale via nationally representative phone surveys during a public health emergency has not been properly validated.

The present study has three objectives. First, to compare the psychometric validity and reliability of ELCSA in a large face-to-face survey conducted prior to the pandemic and three waves of an adapted version collected through a telephone survey. Second, to assess the concurrent validity of the adapted ELCSA scale in the telephone survey. Lastly, to estimate the monthly prevalence of HFI in the general population and among households with and without children during the lockdown of the COVID-19 pandemic in Mexico.

As many other countries struck by the COVID-19, Mexico implemented a national lockdown measure from March 17 to May 31. The Mexican government then shifted to a four-level risk system at the state level. However, during June 2020, most of the country remained at the “red” level, indicating the highest risk, in which only essential businesses could operate and most of the households in the country had to maintain the lockdown.

## Methods

### Data source

ENCOVID-19 is a monthly telephone cross-sectional survey, representative at a national level of individuals 18 years and older who have a mobile phone. It provides information on the well-being of Mexican households during the COVID-19 pandemic. It is publicly available and offers timely information on the social consequences of the lockdown measures in four main domains: labor, income, mental health, and food security. It started in April 2020 and will continue for 12 more months (18). The first ENCOVID-19 survey was collected from April 6 to April 14 (n=833), the second wave was collected between May 20 and May 25 (n=850), and the third wave was collected between June 5 and June 17 (n=1,674).

The monthly surveys were collected based on a one-stage probabilistic sample of mobile telephone numbers which are randomly selected from the publicly available National Dialing Plan (19). The number selection uses a single stratified random sampling for the 32 Mexican states and is implemented with Random Digit Dialing (RDD). As of April 3, 2020, the coverage of mobile phones in Mexico was 96% (19). Previous surveys from the National Institute of Statistics (INEGI for its acronym in Spanish) confirm the wide coverage of mobile phones in Mexico; in 2019, a national survey on the availability and use of technology found the coverage to be 89.4%, but drops to 74% in rural areas (20). Since the ENCOVID-19 might not reach isolated communities, post-stratification sampling weights were used to correct for minor deviations from the Mexican population’s demographic structure. Weights were calculated using the 2015 census data from INEGI and adjust the sample by geographic distribution (state) and by sex, age, and socioeconomic status. Further details of ENCOVID-19 and the composition of the sample are available elsewhere (18).

The monthly surveys were collected by trained interviewers. In addition, a supervisor randomly assessed the quality of interviews though a quality control data management system. On average, the survey was collected in18 minutes using Computer Assisted Telephone Interviewing software (CATI).

### Measures

The study used three cross-sectional ENCOVID-19 waves (i.e., April, May and June). HFI was measured with the 8-item adult version of the ELCSA (14), which is the basis of the Food Insecurity Experience Scale (FIES) (15). ELCSA has been extensively validated for Mexico (21) and is widely used in the country to measure multidimensional poverty (22). It is also included in the Mexican National Health and Nutrition Survey (ENSANUT for its acronym in Spanish). Hence, the last ENSANUT conducted in 2018 was used to compare HFI prevalence before the COVID-19 pandemic.

The ELCSA enquires if, in the last three months, due to a lack of money or other resources, the respondent or any other adult in the household: (i) worried you might run out of food (*worried*); (ii) were unable to eat healthy, balanced, and nutritious food (*healthy*); (iii) ate only a few kinds of foods (*fewfoods*); (iv) skipped breakfast, lunch or dinner (*skipped*); (v) ate less than s/he thought should have (*ateless*); (vi) ran out of food (*ranout*); (vii) were hungry but did not eat (*hungry*); and (viii) went without eating for a whole day (*whlday*). Responses to all items are dichotomous (i.e., Yes/No). Through a total summative score, four levels of food (in)security are estimated: food security (total score =0), mild food insecurity (total score = 1-3), moderate food insecurity (total score = 4-6), and severe food insecurity (total score =7-8). This method of estimation was followed in the current analysis; missing values were minimal, with a maximum of 1.7% in April.

The usual way to use the ELCSA scale is by repeating, for each item, the three-month time-frame and emphasizing lack of money or other resources as the cause to endorse the item. Since telephone surveys need to be short, an adapted version of the ELCSA was used in which the time and lack of resources framing was mentioned only once, before asking the items (see Supplemental Materials S.1). Interviewers were instructed to repeat it whenever the respondent hesitated on the meaning of an item. The 8-item ELCSA was on average collected in four minutes.

Socioeconomic status (SES) was measured with the assets-based AMAI index (23). It combines six household indicators from the National Income and Expenditure Survey (2): (i) education level of the head of household; (ii) number of complete bathrooms; (iv) number of cars or vans; (v) having internet connection; (vi) number of household members 14 years or older who are working; and (vii) number of bedrooms. Based on a summative score and standard cut-off points, SES is categorized into seven mutually exlcusive categories, ranging from “A/B” to “E”, where E represents the lowest SES level.

Anxiety was measured with the the two-item Generalized Anxiety Disorder scale (GAD-2) (24) that inquires about the frequency by which the respondent felt during the last two weeks: nervous, anxious, or on edge; and (ii) not being able to stop or control worrying. Response options are “never”; “several days”; “more than half of days”; and “almost every day”. An additive score of the responses was computed (range 0 to 6), and a cutoff point of 3 or more was used to classify as having anxiety disorder symptoms.

The surveys from May and June included a filter question to identify households with individuals under 18 years; this was unavailable for April. Hence a dummy variable identifying households with children (<18) or without children was generated to address differences in HFI prevalence between these two tpyes of families.

### Analysis

Statistical analyses were conducted in four steps. First, general reliability estimates compared how consistently the ELCSA scale psychometrics performed in the different samples. Alpha and Omega (with tethrachoric correlations) were estimated and compared; according to accepted standards (25) values above 0.7 indicated adequate reliability and suggested high inter-item correlations.

Second, internal psychometric validity was assessed as recommended with the Rasch measurement model (26). The Rasch model shows the extent to which observable items are consistent with the latent phenomenon, i.e. HFI (27). Estimations were conducted with the one-parameter logistic model using a conditional maximum likelihood approach and sampling weights at the household level from each survey (15). Given the eight dichotomous responses to the ELCSA, the models yield item severity parameters indicating the location of the item on the latent variable. Higher severities correspond to items with higher levels of food insecurity and, therefore, with lower item-means. Likewise, item severity was estimated for the total summative score as a way to assess if the items were measuring the whole range of the latent trait.

Item performance was assessed through infit statistics, which show the strength and consistency of the association between an item and the latent trait. The Rasch model assumes all items discriminate equally well and, if the assumption is met, infit statistics equal 1; nonetheless, infit values in the range 0.7-1.3 are considered acceptable (15). When the statistic is between 1.3 and 1.5, items can still be used for measurement but should be closely monitored to establish if there is a systematic bias. Items with infit values above 1.5 should be discarded. Infit values below 0.7 are less worrisome because they indicate the item’s contribution may be redundant.

To assess the comparability of measures of fit between samples a “Flat” reliability test was performed. “Flat” reliability measures the proportion of total variation in true severity accounted for by the model (15). A good model fit has a reliability between 0.7 and 0.8. Lastly, conditional independence of items was examined with an inspection of residual correlations, where it is expected they do not exceed 0.3.

All the analyses were conducted for four surveys: ENSANUT 2018 and three cross-sections of the ENCOVID-19 (i.e., May to June). ENSAUT 2018 served as the pre-pandemic reference (N=44,509). Likewise, the analyses were conducted for households with and without children in the May to June samples.

The third step in the analysis focused in the concurrent validity assessed by correlating the ELCSA score with SES (Spearman correlation) and with anxiety (Welch two-sample test). These analyses were conducted in a pooled dataset of the three months using the raw summative score of food insecurity (range 0-8). We hypothesized that food insecurity would have a negative association with SES and higher levels of anxiety would be found in food insecure households.

In the fourth step we estimated the prevalence of HFI and assessed how much it changed over the first three months of the lockdown. Estimates from ENCOVID-19 were compared with those from ENSANUT 2018. The analysis was performed for the total sample and stratified by households with and without children.

All analyses were conducted in R software; Rasch models were ran using the RM.weights package, developed by FAO to conduct the statistical validation of the FIES (28).

## Results

The adapted version of the ELCSA scale used in ENCOVID-19 repeated cross-sectional surveys was found to be reliable. The Alpha coefficient for ENSANUT was 0.90 and the values for ENCOVID-19 –April, May and June– ranged between 0.87 and 0.89. A more stringent measure for dichotomous matrices showed similar results. While ENSANUT had an Omega coefficient of 0.80, the values of the ENCOVID were between 0.75 and 0.77. Inter-item correlations were above the cut-off point of 0.6 in all surveys.

ELCSA’s item severity parameters in the four surveys followed a similar order as the FIES global standard (15) (Table 1). Severity parameters inversely mirror the mean’s order and showed how the adapted version of the ELCSA in the ENCOVID-19 was able to measure the full range of the latent trait. ELCSA item’s psychometric performance was adequate in the ENCOVID-19 surveys (Table 1). The only item with an infit statistic above the 1.3 threshold was “ran out of food” (*ranout)*, in May’s wave, with a value of 1.35. This was not a concern since it represented a small deviation from the range, it had the highest infit value in the ENSANUT survey (1.24) and the “misfit” was not systematic. No item had systematic deviations in the outfit values (Table 1).

**Table 1.**
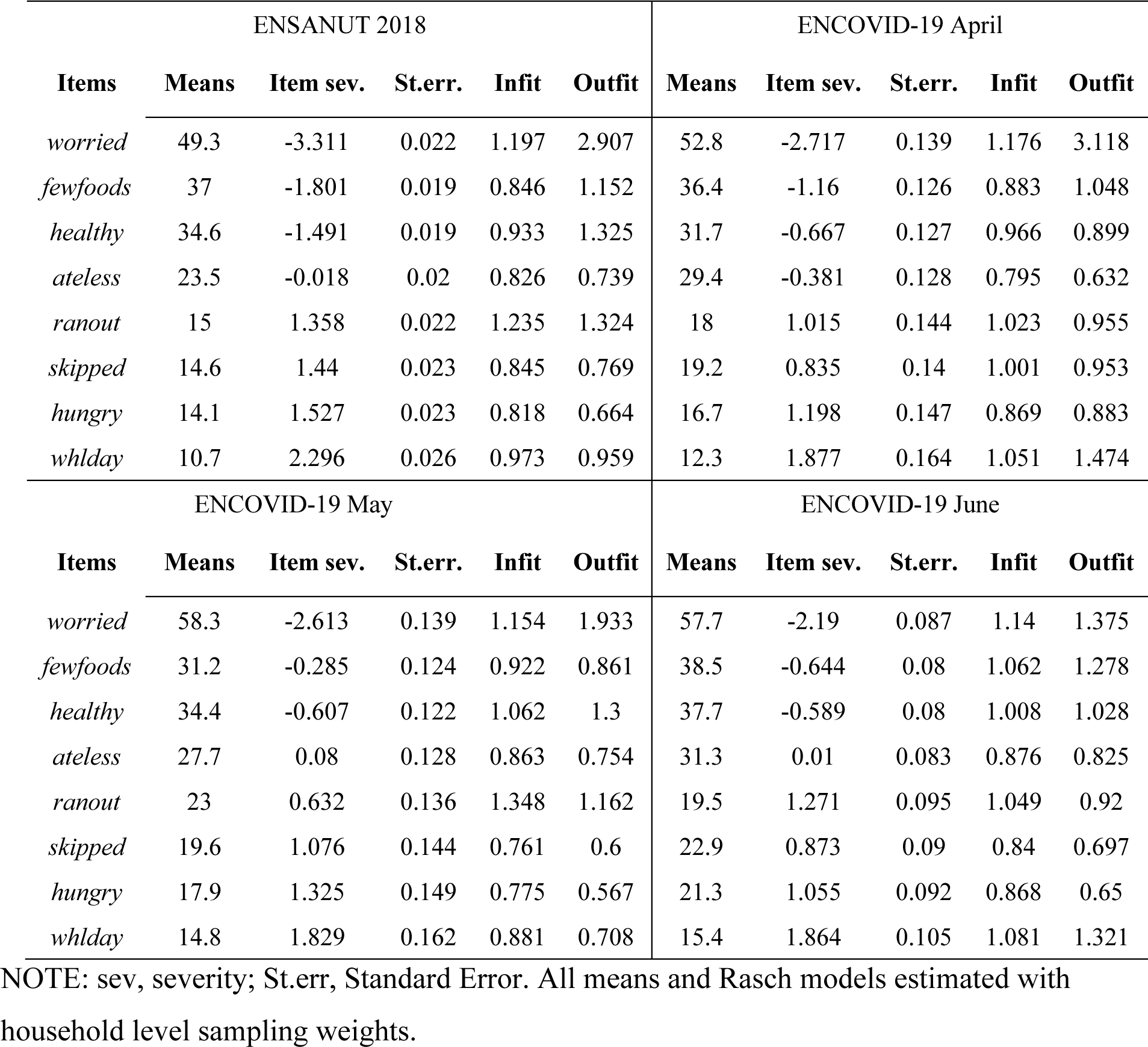
Comparison of weighted means and item severity parameters, infit and outfit statistics of the ELCSA scale between the ENSANUT 2018 and the three ENCOVID-19 surveys from April, May, and June 2020

The severity parameters of the raw summative score were equivalent between surveys (Table 2). These results indicate that the summative score of ELCSA from ENCOVID-19 was able to measure equally well the three levels of HFI when compared with ENSANUT 2018. Overall model fit was excellent in the ENCOVID-19 surveys. Flat reliability had a value of 0.78 in the ENSANUT survey and was very similar in ENCOVID-19, April (0.74), May (0.73) and June (0.73). The assumption of conditional independence held for most pairs of items. The internal validity in ENCOVID-19 in households with and without children in May and June also confirmed its adequate psychometric performance (see Supplementary Materials S.2).

**Table 2.** Comparison of the severity parameters from the raw summative score of the ELCSA scale between the ENSANUT 2018 and the three ENCOVID-19 surveys from April, May, and June 2020

| Score | ENSANUT 2018 |  | ENCOVID-19<br>April |  | ENCOVID-19<br>May |  | ENCOVID-19<br>June |  |
| --- | --- | --- | --- | --- | --- | --- | --- | --- |
|  | Severity | St.err. | Severity | St.err. | Severity | St.err. | Severity | St.err. |
| 0 | -4.106 | 1.601 | -3.589 | 1.577 | -3.363 | 1.601 | -3.187 | 1.552 |
| 1 | -3.076 | 1.252 | -2.654 | 1.218 | -2.386 | 1.239 | -2.273 | 1.180 |
| 2 | -1.779 | 1.058 | -1.500 | 0.973 | -1.213 | 0.966 | -1.209 | 0.941 |
| 3 | -0.875 | 1.001 | -0.656 | 0.877 | -0.399 | 0.850 | -0.429 | 0.848 |
| 4 | 0.178 | 0.942 | 0.080 | 0.842 | 0.275 | 0.809 | 0.254 | 0.814 |
| 5 | 1.026 | 0.907 | 0.788 | 0.846 | 0.933 | 0.817 | 0.936 | 0.823 |
| 6 | 1.815 | 0.935 | 1.544 | 0.908 | 1.648 | 0.887 | 1.631 | 0.888 |
| 7 | 2.889 | 1.144 | 2.539 | 1.132 | 2.613 | 1.120 | 2.624 | 1.123 |
| 8 | 3.736 | 1.601 | 3.368 | 1.577 | 3.429 | 1.601 | 3.437 | 1.552 |

NOTE: St.err, Standard Error. Rasch models estimated with household level sampling weights.

Concurrent validity of food insecurity was established by its association with SES and anxiety. The Spearman correlation between the raw score of food insecurity and SES was negative, and statistically significant (−0.4; CI: -0.37—0.43). Figure 1 shows a clear dose-response gradient between SES and HFI severity. Even though mild HFI was present at every SES level, moderate and severe food insecurity increased as SES dropped. At the lowest SES level, moderate and severe food insecurity reached its highest prevalence, at 28.9% and 20.9%, respectively. Notably, middle-SES households reported a prevalence of moderate and severe food insecurity from 10% in C+ to 26% in D+.

**Figure 1.**
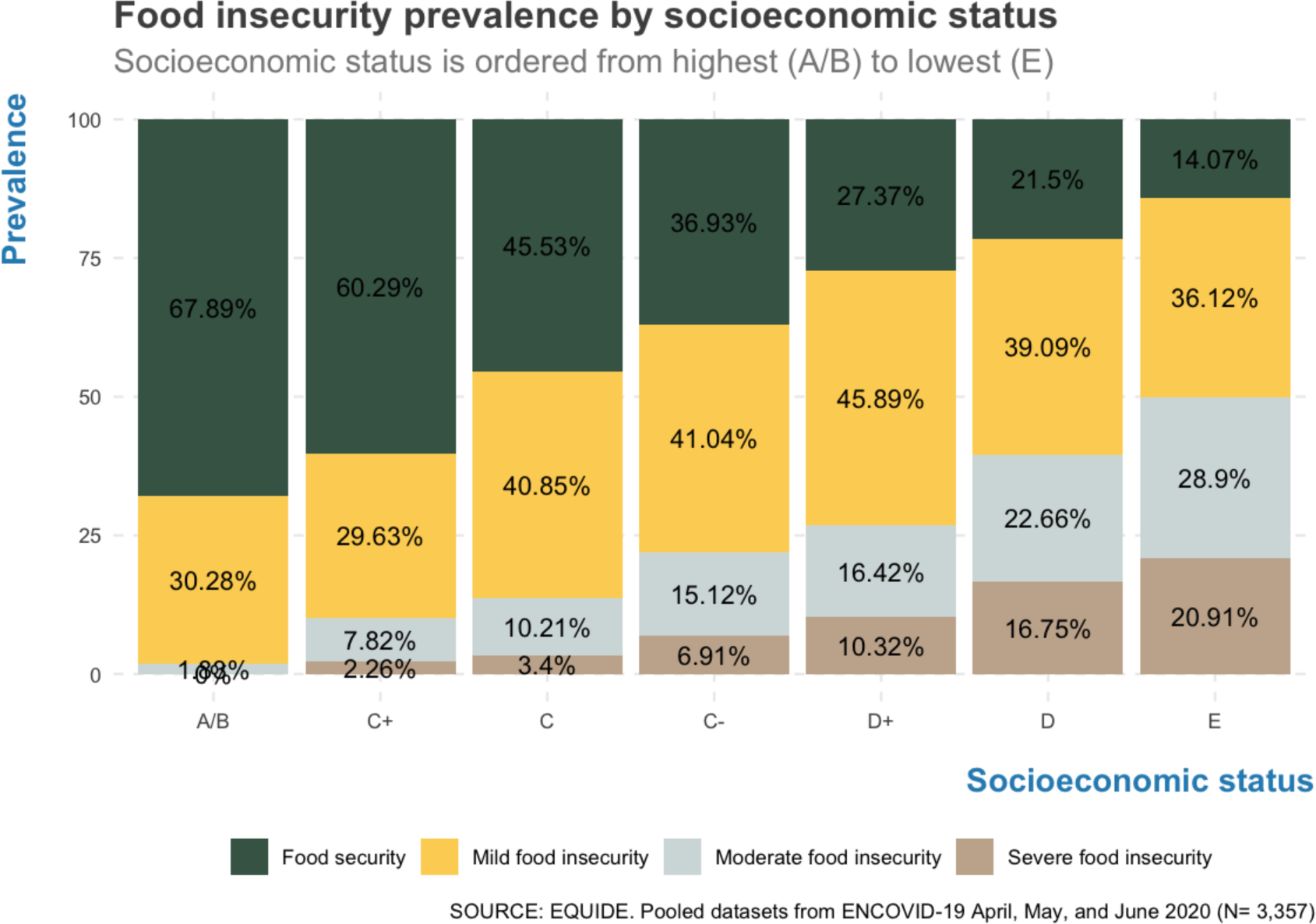
Food insecurity prevalence by socioeconomic status. The graph illustrates the inverse relationship between socioeconomic status and food security. Households in the lowest SES level have the highest prevalence of moderate and severe food insecurity.

Anxiety was also associated with HFI in the expected direction. Individuals without anxiety symptoms reported a mean total food insecurity score of 1.68 and this score was substantially higher among individuals with anxiety symptoms (3.11). Figure 2 shows the dose-response gradient between HFI and anxiety. While 19.3% of persons living in food secure households reported symptoms of anxiety, this was the case for 57.1% among those living in households with severe food insecurity.

**Figure 2.**
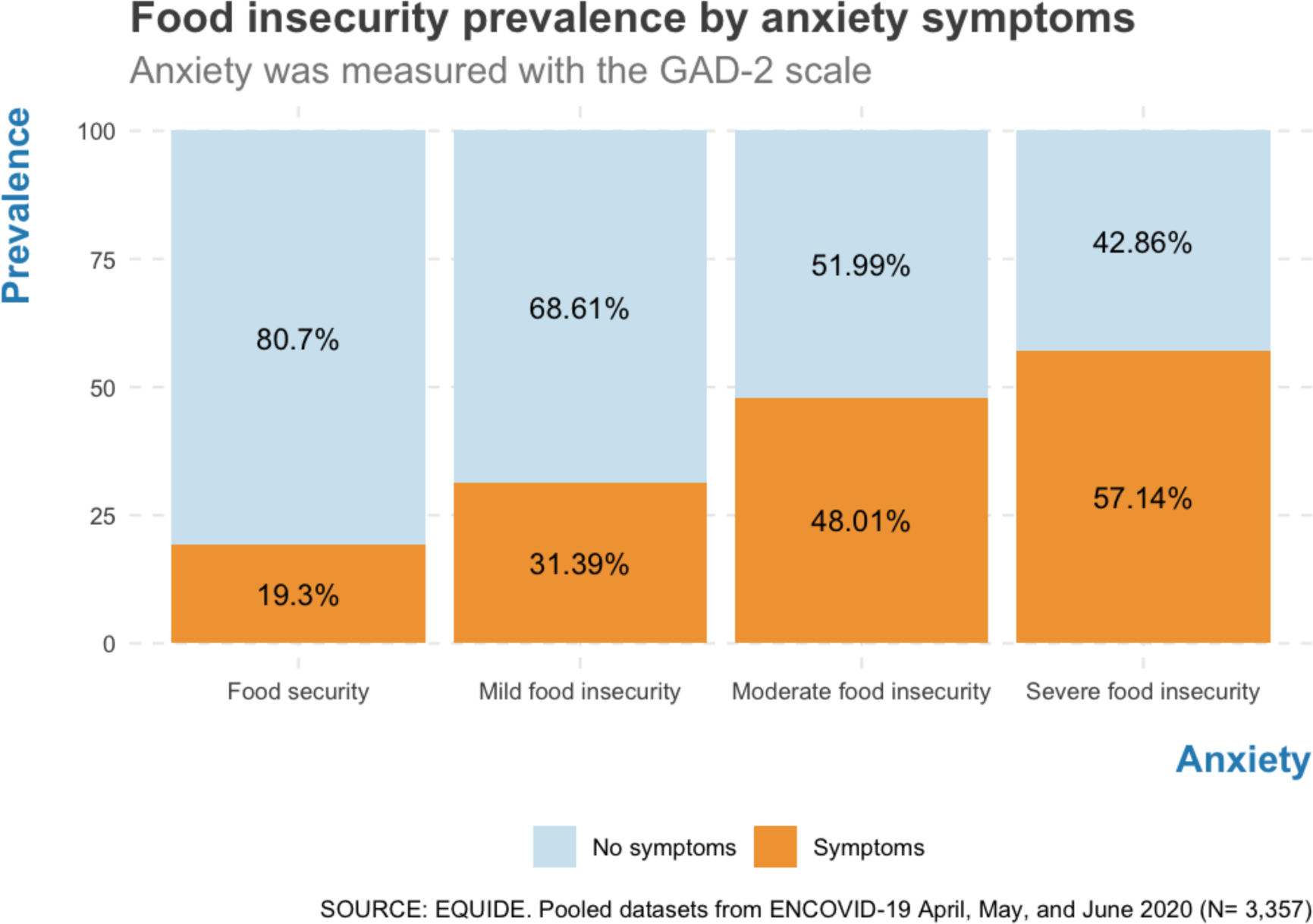
Food insecurity prevalence by anxiety symptoms. The graph illustrates the positive relationship between socioeconomic status and anxiety. Persons living in food secure households report fewer anxiety symptoms than persons living in food insecure households

Prevalence estimates indicated that the prevalence of food security has been decreasing as the COVID-19 pandemic advances in Mexico (Table 3). According to ENSANUT 2018, 44.7% of households were food secure, but then it significantly dropped to 38.8% in April, and then to 33.2% in May, and 30.6% in June, 2020 (Figure 3). Mild HFI reached its highest level in May (41.7%) and moderate food insecurity in June, 2020 (18.65%). Severe food insecurity in June was not statistically different from the 2018 prevalence, however, the secular trend suggested that severe food insecurity may also be worsening on a monthly basis.

**Table 3.** Prevalence comparisons between the ENSANUT 2018 and the three ENCOVID-19 surveys from April, May, and June 2020

|  | ENSANUT 2018 |  |  | ENCOVID-19 April |  |  | ENCOVID-19 May |  |  | ENCOVID-19 June |  |  |
| --- | --- | --- | --- | --- | --- | --- | --- | --- | --- | --- | --- | --- |
|  | Prop | CI |  | Prop | CI |  | Prop | CI |  | Prop | CI |  |
| **Food Security** | 44.75 | 43.93 | 45.57 | 38.88 | 35.59 | 42.18 | 33.19 | 29.64 | 36.74 | 30.6 | 28.02 | 33.18 |
| **Mild FI** | 31.09 | 30.43 | 31.75 | 33.6 | 30.33 | 36.87 | 41.73 | 37.73 | 45.72 | 39.22 | 36.41 | 42.03 |
| **Moderate FI** | 14.9 | 14.36 | 15.43 | 17.18 | 14.53 | 19.82 | 13.31 | 10.33 | 16.29 | 18.65 | 16.42 | 20.89 |
| **Severe FI** | 9.26 | 8.84 | 9.68 | 10.33 | 8.2 | 12.45 | 11.75 | 8.84 | 14.66 | 11.51 | 9.57 | 13.45 |

NOTE: All proportions were estimated with household level sampling weights. Prop, Proportion; CI, Confidence interval.

**Figure 3.**
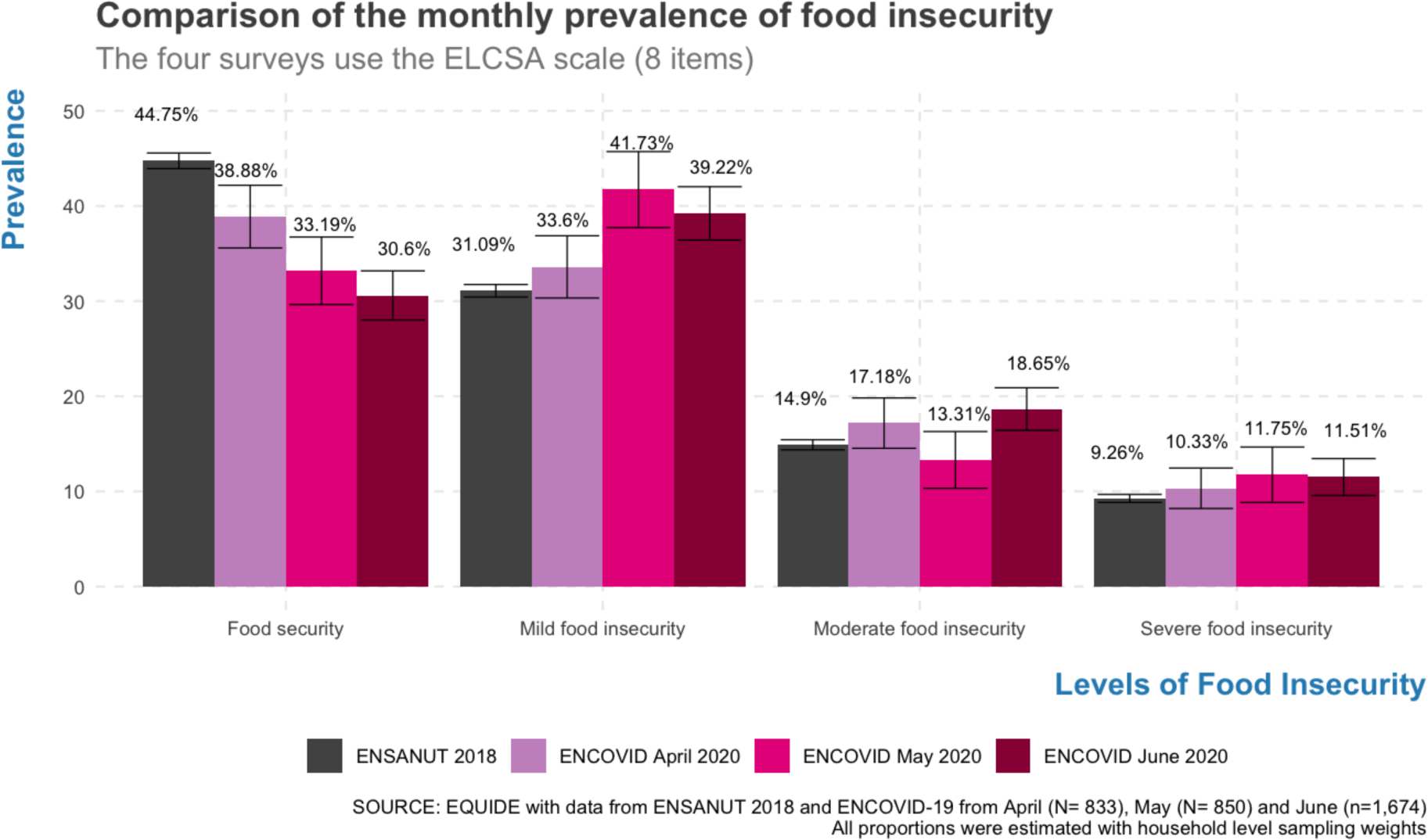
Monthly comparison of the prevalence of food insecurity. In contrast with ENSANUT from the year 2018, food security is decreasing during the COVID-19 pandemic. Mild food insecurity reached its highest level in May and moderate food insecurity was highest in June.

**Figure 4.**
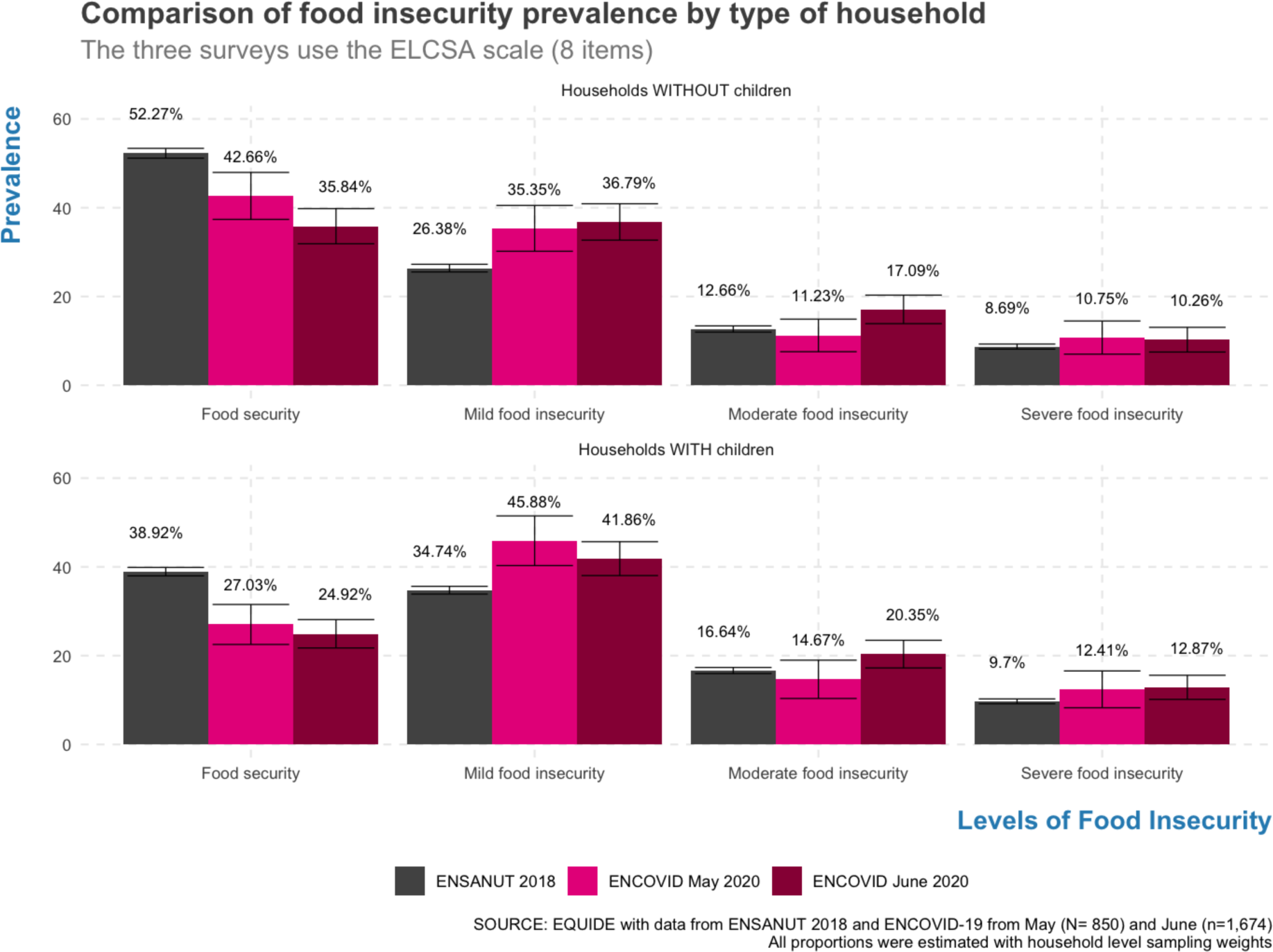
Comparison of the prevalence of food insecurity by type of household. Household with children have lower levels of food security than households without children. Households with children had the highest prevalence of mild food insecurity in May and of moderate food insecurity in June 2020.

The stratified analysis showed that HFI was higher in households with than without children. Food security in households with children decreased from 38.9% in 2008 to 27% in May and to 24.9% in June. The highest prevalence of mild food insecurity was during May (45.8%) and for moderate food insecurity during June (20.3%). Severe food insecurity seems to be increasing as well but confidence intervals overlapped with those of ENSANUT 2018 (see Supplementary Material S.2).

## Discussion

Our findings strongly suggest that the COVID-19 pandemic is increasing HFI in Mexico during the mandated lockdown. These results are consistent with previous studies that demonstrate how vulnerable household food security is during socioeconomic shocks (5). Moreover, prevalence estimates showed that food security was decreasing on a monthly basis. Furthermore, long lockdowns associated with the pandemic might keep disrupting food systems and food insecurity can worsen even more as a result (3).

The adapted version of the ELCSA scale used in the ENCOVID-19 proved to be reliable and valid. Rasch models showed that the high reliability of the telephone version of the scale was comparable to the face-to-face application in ENSANUT 2018. A limitation of ENCOVID-19, however, was the insufficient inclusion of persons living in rural and isolated localities due to lower mobile phone coverage. Since these communities have higher extreme poverty levels than urban enclaves in Mexico (29), lower coverage of these areas may have led to an underestimation of HFI. In spite of this limitations, thanks to ENCOVID-19, Mexico has been able to rapidly and efficiently monitor changes in food security in the general population and diverse vulnerable locations. A necessary next step is thus to further study the determinants of HFI during and post-pandemic in low-income households that are being affected the most, such as single women headed household or caregivers working in the informal sector. Analyzing these determinants will be key to designing adequate public policies.

The validity of ELCSA was also examined in its association with other measures. Households with low SES reported a considerably higher prevalence of food insecurity than those with high SES. Remarkably, food insecurity was reported in 10% to 26% of households in the middle of the SES scale. This means that the pandemic is striking households along the SES continuum, but it is hitting harder the ones with lower SES.

HFI was also associated with anxiety. Individuals living under conditions of moderate and severe HFI had more than double the anxiety levels than food secure households. These results confirm the expected psycho-emotional toll of experiencing HFI during the pandemic. It is possible that the increase in food insecurity in households with young children may be leading to more family chaos and poor interpersonal relations (30). Likewise, the stress levels in households with food insecurity may have increased the odds of suffering emotional and physical intimate partner violence (31). It is important to keep monitoring the interaction of the two indicators because these consequences will most likely intensify with the additional stress of the economic and health crisis. Examining these associations over time can indeed shed light on the syndemic effects of food insecurity on household dynamics during the pandemic (3), and may help understand ways of intervining to address the reinforcing association between anxiety and HFI.

Food insecurity is more prevalent in households with children. These households require focalized social protection actions. The definition of children in ENCOVID-19 is not as granular as would have been desirable, as clusters of infants, children and adolescents, which require different policy strategies. Prior studies have already highlighted the vulnerability of young children (32) during the COVID-19 pandemic. The findings of the worsening trend of food insecurity stress the urgent need for policy actions. It is important to consider providing and evaluating food assistance and cash transfers, at least to the most vulnerable households. For instance, prior studies have found that a cash transfer of one minimum wage and a waiver of payment of basic services (i.e. electricity, water, etc.) to households with informal jobs and with children under 5 years of age may help protect their food security and it would require using less than 0.06% of Mexico’s GDP (33). Moreover, it is important to adapt and continue food assistance programs –especially those in schools when they reopen– directed to quarantined families with children, adolescents, pregnant and lactating women (3). Beyond specific programs, tackling these convergent syndemic-like crises will require multi-level and evidence-based policies based on a complex adaptive systems framework (2). Even if the COVID-19 recedes, its socioeconomic and health effects are expected to last for a long time.

Timely and high-quality HFI surveillance systems can help governments respond to the major food security challenge posed by the COVID-19 pandemic. Telephone surveys are a feasible and cost-effective strategy to measure food insecurity with experience-based scales such as ELCSA. These results can inform similar strategies in countries using the FIES scale, but they might be especially useful for countries in Latin America and the Caribbean that regularly measure food insecurity with the ELCSA scale. ENCOVID-19 is an efficient strategy to regularly monitor changes in several indicators, including HFI, and thus can serve as a high quality instrument that can be frequently applied to inform decision-makers and assess the performance of social protection programs during public health emergencies and other pressing circumstances (34).

## Conclusion

HFI is worsening as the COVID-19 pandemic advances in Mexico. The causal web between the socioeconomic shocks, food insecurity and adverse health and mental health outcomes needs to be addressed as a complex syndemic with comprehensive and multi-level policy actions. Governments should not attempt to navigate the COVID-19 pandemic blindfolded. High-quality and cost-effective strategies to monitor food insecurity are available and should be implemented widely.

## Data Availability

Data is available for April in ZENODO as: Encuesta Nacional sobre los Efectos del COVID-19 en el Bienestar de los Hogares Mexicanos (ENCOVID-19-ABRIL). In a few weeks the other datasets will be released.

http://doi.org/10.5281/zenodo.3950528

